# Attention Deficit and Hyperactivity Disorder and Enuresis co-occurrence in the pediatric population: a systematic review and meta-analysis

**DOI:** 10.1101/2021.01.23.21250367

**Authors:** Ana Cecília de Sena Oliveira, Bruno da Silva Athanasio, Flávia Cristina de Carvalho Mrad, Monica Maria de Almeida Vasconcelos, Maicon Albuquerque Rodrigues, Débora Marques de Miranda, Ana Cristina Simões e Silva

## Abstract

**Background:** Attention Deficit and Hyperactivity/Impulsivity Disorder (ADHD) and Enuresis are common behavioral disorders in childhood, impacting in adolescence and adult life.

**Objective:** We systematically search the literature to verify the relationship between ADHD and enuresis and how these conditions can modify each other during development.

**Method:** Using PRISMA guidelines, we tried to answer the following question: How frequent is ADHD and Enuresis comorbidity?

**Results:** Twenty-five studies were fully read and show similar rates of ADHD in the Enuretic group than the frequency of Enuresis in the ADHD group. There is a two-times higher risk to have both conditions simultaneously.

**Conclusion:** Enuresis and ADHD seems to happen as a continuous of the same spectrum. Further studies are necessary to evaluate if gender, age, course and presence of comorbidities are similar in patients with both conditions in comparison to those with only one of these conditions.

## 1. Introduction

Attention Deficit and Hyperactivity/Impulsivity Disorder (ADHD) is the most common neurodevelopmental disorder reaching up to 6% of children worldwide (Polanczyk et al., 2007). ADHD is a behavioral disorder that can be classified into three subtypes: Inattentive (ADHD-I), Hyperactive-Impulsive (HI) and Combined (ADHD-C). Comorbidities are more a rule than an exception for ADHD, some of them seem to share common pathophysiological mechanisms and implicate in more functional compromise (Weissenberger et al., 2017). For sharing the same origins sometimes the comorbidities appear along life complicating the treatment and the outcomes, while other ones disappear as the children grow and the symptoms fade.

Enuresis is a frequent behavior observed in ADHD children (Baeyens, Roeyers, D’Haese, et al., 2007; Courtney, 2011); (Biederman et al., 1995). Enuresis is characterized by the lack of control of the urinary bladder that results in repeated episodes of involuntary micturition during sleep (day or night) (Baeyens, Roeyers, D’Haese, et al., 2007); (Mota et al., 2015). According to the Diagnostic and Statistical Manual of Mental Disorders (DSM V) (“Anxiety Disorders,” 2013), the Enuresis diagnosis consists of bedwetting frequency of twice per week for three consecutive months or the presence of clinically significant distress or impairment, irrespective of the age of the child (DSM V)(“Anxiety Disorders,” 2013). Enuresis can be classified as primary or secondary. Primary enuresis occurs when individuals have never been able to control micturition or have a previous dry period less than six months (Nevéus et al., 2020). Primary enuresis seems to have a genetic component, the incidence of enuresis is about 15% if none of the parents or their immediate relatives suffered from enuresis, about 44%, if one of the parents or their immediate relatives suffered from enuresis and can be up to 77% if both parents have a positive story (Bogaert et al., 2020; Butler et al., 2005). Secondary enuresis refers to children who have previously experienced at least six months of dryness, but who have experienced relapses. Another essential clinical subdivision is monosymptomatic and non-monosymptomatic enuresis, the last term reserved for the children who besides enuresis have others lower urinary tract symptoms (as daytime incontinence, urgency, voiding difficulties, abnormal voiding frequency) (Nevéus et al., 2020). However, Biederman et al. (1994) found that enuresis does not seem to be modified by primary versus secondary.

ADHD and enuresis are common comorbidities, both associated with neurological maturational delay and potential changes during the brain development (Plummer et al., 2016). Biederman et al. (1994) believe that enuresis does not increase the risk for psychopathology or psychosocial adversity or developmental immaturity. However, the authors pointed out that enuresis is associated with increased risk for learning disability, intellectual deficiency, and worse school achievement in typical development children, but not in children with ADHD (Biederman et al., 1995). Besides the maturational issue, other factors are genetic background, sleep disorders, anatomic, behavioral and psychological conditions, and reduced secretion of antidiuretic hormone (Biederman et al., 1995). An insightful commentary published in 1999 suggests more potential causes for enuresis beyond the inattentive and impulsive behaviors (Järvelin, 1999). However, up to now, there is little information about development, comorbidities, prenatal and perinatal events in most studies.

The relationship between ADHD and nocturnal enuresis has instigated too much discussion, but the shared mechanisms and genetic features still to be known. Both conditions seem to be associated with neurological maturational delay. Children with concomitant ADHD and Enuresis use more therapies than children with enuresis-only, and children with ADHD tend to persist with enuresis for two years of treatment (Baeyens, Roeyers, D’Haese, et al., 2007; Baeyens, Roeyers, Van Erdeghem, et al., 2007). Of the pharmacologic approaches, the interventions have some common medications that can help us to understand the relationship between these two disorders (Fritz et al., 2004). There is also evidence suggesting that Desmopressin (DDAVP) produces arousing properties that might involve central dopaminergic mechanisms (Di Michele et al., 1996).

In this review, we systematically search the literature to verify the relationship between ADHD and enuresis and how these conditions can modify each other during development. Using PRISMA guidelines, we tried to answer the following question: How frequent is ADHD and Enuresis comorbidity?

## 2. Objective

We aimed to answer the following question: How frequent is ADHD and Enuresis comorbidity? Secondary questions emerged with the review about sociodemographic characteristics and comorbidities of populations with Enuresis and ADHD.

## 3. Methods

### 3.1. Design

This systematic review was conducted according to the guidelines of the Preferred Reporting Items for Systematic Reviews and Meta-Analyses (PRISMA) (Moher et al., 2009). PROSPERO register number is CRD42020208299.

### 3.2. Eligibility criteria

We considered eligible for inclusion case control, cohort, and cross sectional studies examining pediatric population on the co-occurrence of ADHD and enuresis. ADHD patients had to be diagnosed according to DSM or ICD criteria. Studies addressing enuresis, but who did not include patients with nocturnal enuresis, only daytime incontinence, were excluded.

The following articles were also excluded from this review: (i) articles other than the specified in inclusion criteria: case report, case series, clinical trials and review articles; (ii) articles that were not written in the English language; (iii) articles that did not specify diagnostic criteria for both ADHD and enuresis; (iv) papers addressing neurologic or psychiatric disorder other than ADHD.

### 3.3 Search methods for identification of studies

Search was carried out by two independent authors, BSA and ACSO, in three databases: PUBMED, Scopus and SciELO. Articles published until June 2020 were included in this review. No language restrictions were applied. The search terms utilized were “ADHD” and “enuresis”. The search combinations used was: ((ADHD) OR (Attention Deficit and Hyperactivity Disorder)) AND (enuresis).

Duplicates were excluded and studies were initially extracted for abstract screening. Those found relevant were retrieved for full text read. Whenever necessary, further data were requested to authors. Disagreements on eligibility were resolved in discussions between authors who extracted data from studies defined eligible.

To address potential bias, studies were assessed for level of evidence. Case control and cohort studies were evaluated according to the Ottawa-Newcastle Scale (NOS) (Lo et al., 2014). For cross sectional studies, authors addressed potential bias in the Discussion section.

### 3.4 Meta-analysis

Prevalence data was merged in a meta-analysis about the pooled odds ratio when studies have the same design and comparable populations. Two strategies were adopted. First, there is a combined analysis of studies including patients with and without ADHD informing the frequency of enuresis. Second, pooled evaluation of studies including Enuretic and Non-Enuretic patients in which ADHD frequency was investigated.

The odds ratio (OR) was used as a summary statistic test (Chang and Hoaglin, 2017). I^2^ test was used to evaluate the heterogeneity and to assess between-study inconsistency, I^2^ statistic was used and classified as low (I^2^=25%), medium (I^2^=50%), and high (I^2^=75%) heterogeneity (Huedo-Medina et al., 2006). χ^2^ p-value lower than .1 or I^2^ higher than 50% indicated the presence of heterogeneity among studies, so the random effects model was used for analysis. Otherwise, the fixed effects model was applied (Feng et al., 2017). Every analysis was performed using RevMan 5.4.0 software.

## 4. Results

Among the 485 screening articles the exclusion criteria for study type were: clinical trials and case reports, case series and reviews, letters, and editorials. After eligibility, 25 studies were selected for qualitative synthesis included case-control studies (n=9), cohort studies (n=3) and cross sectional studies (n = 13).

In order to address potential bias, case control and cohort studies were assessed for the level of evidence with the Ottawa-Newcastle Scale (NOS) (Lo et al., 2014), as shown in Tables 1 and 2. Limitations and risks of bias in cross sectional studies were also considered.

**Table 1.** Assessment of the studies by the Newcastle - Ottawa Scale (NOS) quality assessment scale case control studies

| Authors | Selection | Comparability | Exposure |
| --- | --- | --- | --- |
| (Zavadenko et al., 2011) |  | ** | * |
| (Rodopman-Arman et al., 2011) | **** | ** | ** |
| (Okur et al., 2012) | *** | ** | ** |
| (Baeyens, Roeyers, D'Haese, et al., 2007) | **** | ** | ** |
| (Robson et al., 1997) | *** | ** | ** |
| (J. Niemczyk et al., 2015) | *** | ** | ** |
| (Yousefichaijan, P., Sharafkhah, M., Salehi, B., Rafiei, M, 2016) | *** | ** | ** |
| (Biederman et al., 1995) | * | ** | ** |
| (Abd-Elmoneim et al., 2020) | ** | ** | ** |

**Table 2.** Assessment of the studies by the Newcastle - Ottawa Scale (NOS) quality assessment scale cohort studies

| Authors | Selection | Comparability | Outcome |
| --- | --- | --- | --- |
| (Mellon et al., 2013) | **** | ** | *** |
| (Cak et al., 2013) | ** | ** | *** |
| (Kessel et al., 2017) | **** | * | *** |
| (Baeyens, Roeyers, Van Erdeghem, et al., 2007) | **** | ** | ** |

### 4.1) Studies characteristics and methodological issues

Selected papers differed in study type, research question and controls utilized. For clarity in qualitative synthesis, articles were separated according to subjects and controls as follows: studies with ADHD subjects screened for enuresis (n= 7, Table 3); studies with enuresis subjects screened for ADHD (n=5, Table 4); studies that screened ADHD and non-ADHD subjects for NE (n=7, Table 5); studies that screened enuresis and non-enuresis subjects for ADHD (n=6, Table 6). Whenever necessary, differences in study designs within these categories were highlighted.

**Table 3.** Studies that screened attention deficit hyperactivity disorder (ADHD) patients for enuresis (E)

| Author | Enuresis criteria | ADHD criteria | Sample Size (NE/non-NE) | Mean age ± SD and/or age range (years) | E % (E/total) | Gender (Male %) | | | |
| --- | --- | --- | --- | --- | --- | --- | --- | --- | --- |
|  |  |  |  |  |  | ADHD | ADHD + E | ADHD + non-E | p-value |
| (Zavadenko et al., 2011)** | ICD-10 | ICD-10 | 53/71 | 5-14yrs (5-9 and 10-14yrs) | 53/124 = 42,7% | 83,30 % | 73.5% | 80.2% | n.s. |
| (Ghanizadeh, 2010) ** | DSM-IV | KSADS-IVR (DSM-IV) | 35/153 | 9.2 ± 2.6yrs | 35/188 = 18,6% | 77.1% | 85.7% | 75.1% | p = 0.10 |
| (Elia et al., 2009) | CBCL | KSADS-IVR (DSM-IV) | 58/286 | E: 8.23 ± 1.6yrs<br>non-enuresis : 8.6 ± 1.7<br>p= 0,15 | 58/314 = 18,4% | 76.4% | 82.7% | 75.1% | p=0.24 |
| (Yang et al., 2013) | DVSS | DSM-IV | 15/38 | E: 7.53 ± 1.06yrs<br>non NE: 7.26 ± 1.03<br>p = 0,40 | 15/53 = 28,3% | 88.6% | 100% | 84.2% | p=0.10 |
| (Amiri et al., 2013) | CTRS | DSM - IV | 28/132 | 9.26 ± 1.14 yrs | 28/160 = 17,5% | 52.9% | 60.7% | 63.4% | n.s. |
| (Khazaie et al., 2018) | Own methods* | DSM-IV-TR | 111/220 | 7.9 ± 1.3yrs | 111/331 = 33,5% | 61,30 % | 72.0% | 56.8% | p=0.007 |
| (Ghanizadeh, 2009)** | DSM-IV | DSM-IV | 20/171 | 9.0 ± 2.5yrs | 20/191 = 10,4% | 79.5% | 95.0% | 77.3% | p=0.07 |

\*Pediatric urologist and urinalysis, renal sonography, and interview with children and their parents.
\*\* Enuresis
ICD-10 = International Classification of Diseases, Tenth Revision
DSM = Diagnostic and Statistical Manual of Mental Disorders 4th Edition
CTRS = Conners' Teacher Rating Scale
DVSS = Dysfunctional Voiding Scoring System

**Table 4.**
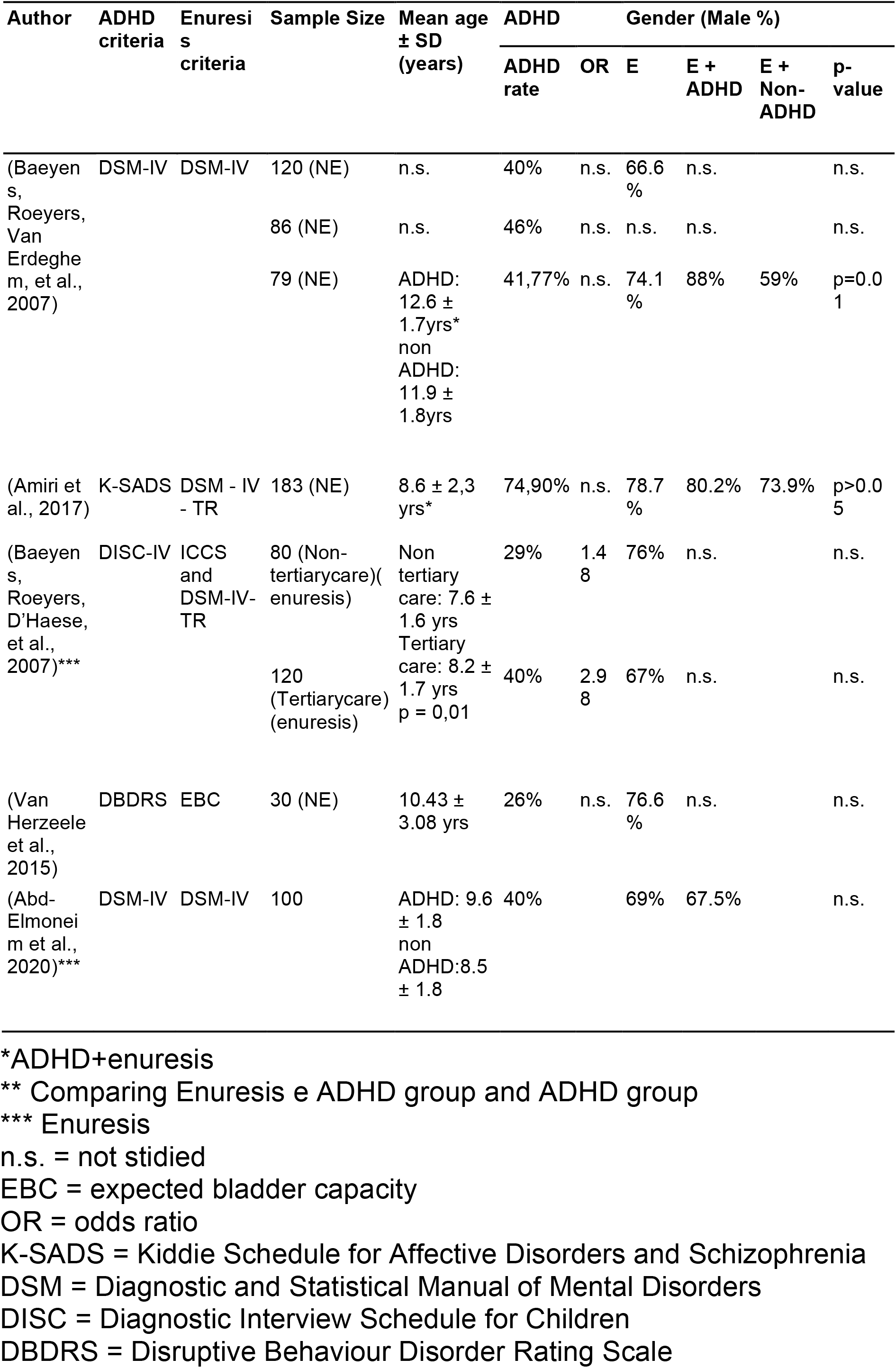
Studies that screened patients with enuresis (E) for attention deficit hyperactivity disorder (ADHD)

**Table 5.** Studies that screened attention deficit hyperactivity disorder (ADHD) and non-ADHD patients for enuresis (E)

| Author | Enuresis criteria | ADHD criteria | Sample Size (ADHD/Controls) | ADHD age (mean ± SD) | E | | | Gender (Male %) | | |
| --- | --- | --- | --- | --- | --- | --- | --- | --- | --- | --- |
|  |  |  |  |  | E in ADHD group | E in non-ADHD group | p-value | E + ADHD | E + Non-ADHD | p-value |
| (Rodopman-Arman et al., 2011) | CSHQ | K-SADS | 44/40 | ADHD: 9.1 ± 1.5 yrs<br>Control s: 9.3 ± 1.3<br>p = 0,24 | 15.1 % (7/44) | 3.3 % (1/40) | p = 0.001 | 80% | 75% | p=0.36 |
| (Robson et al., 1997) | o.q. | DSM-III R | 153/142 | ADHD: 11.2 ± 1.9 yrs<br>Control s: 10,2 ± 2,4 yrs<br>p<0,01 | 20.9 %** (32/153) | 7.7 % (11/142) | p = 0.05 | 57% | 43% | p=0.02 |
| (J. Niemczyk et al., 2015) | ICCS | DSM-IV | 40/43 | 11.4 ± 2.49 yrs | 5 % (2/40) | n.s. | n.s. | 75% | 60.5% | n.s. |
| (Biederman et al., 1995)*** | DSM-III-R | DSM-III-R | 140/120 | 10.1 ± 2.7 yrs* | 32.14% (45/140) | 14.16% (17/120) | p < 0.001 | 100% | 100% | n.s. |
| (Mellon et al., 2013)*** | DSM-IV | DSM-IV | 358/729 | ADHD: 6.7 ± 2.1 yrs<br>Control s: 7.0 ± 2.5 yrs | 7 yrs: 9.7 % (35/358)<br>15 yrs: 9,9% (3.4/35) | 7 yrs: 4.6% (34/729)<br>15 yrs: 4.8%(1.6/34) | p = 0.002 | 74,50% | 75,20 % | n.s. |
| (Cak et al., 2013) | K-SADS-PL | DSM-III R | 45/28 | 14.17 ± 1.04 yrs (7-10 and 13-16) | 7-10: 26.7 % (12/44)<br>13-16: 0% | 7-10: 7.1 % (2/28)<br>13-16: 0% | n.s. | 71.1% | 75% | n.s. |
| (Tsai et al., 2017) | DSM-IV | DSM-IV | 14900/59600 | 10 ± 2.49 yrs | 1.15% (171/14900) | 0.48% (286/59600) | p < 0,001 | 79.4% | 79.4% | p=0.99 |

\*ADHD + enuresis
\*\* When subjects were 6-years-old
\*\*\* Enuresis
o.q. = own questionnaire
CSHQ = Children's Sleep Habits Questionnaire
ICCS = International Children's Continence Society
K-SADS = Kiddie Schedule for Affective Disorders and Schizophrenia
KSADS-PL = KSADS-Present and Lifetime Version
DSM = Diagnostic and Statistical Manual of Mental Disorders

**Table 6.**
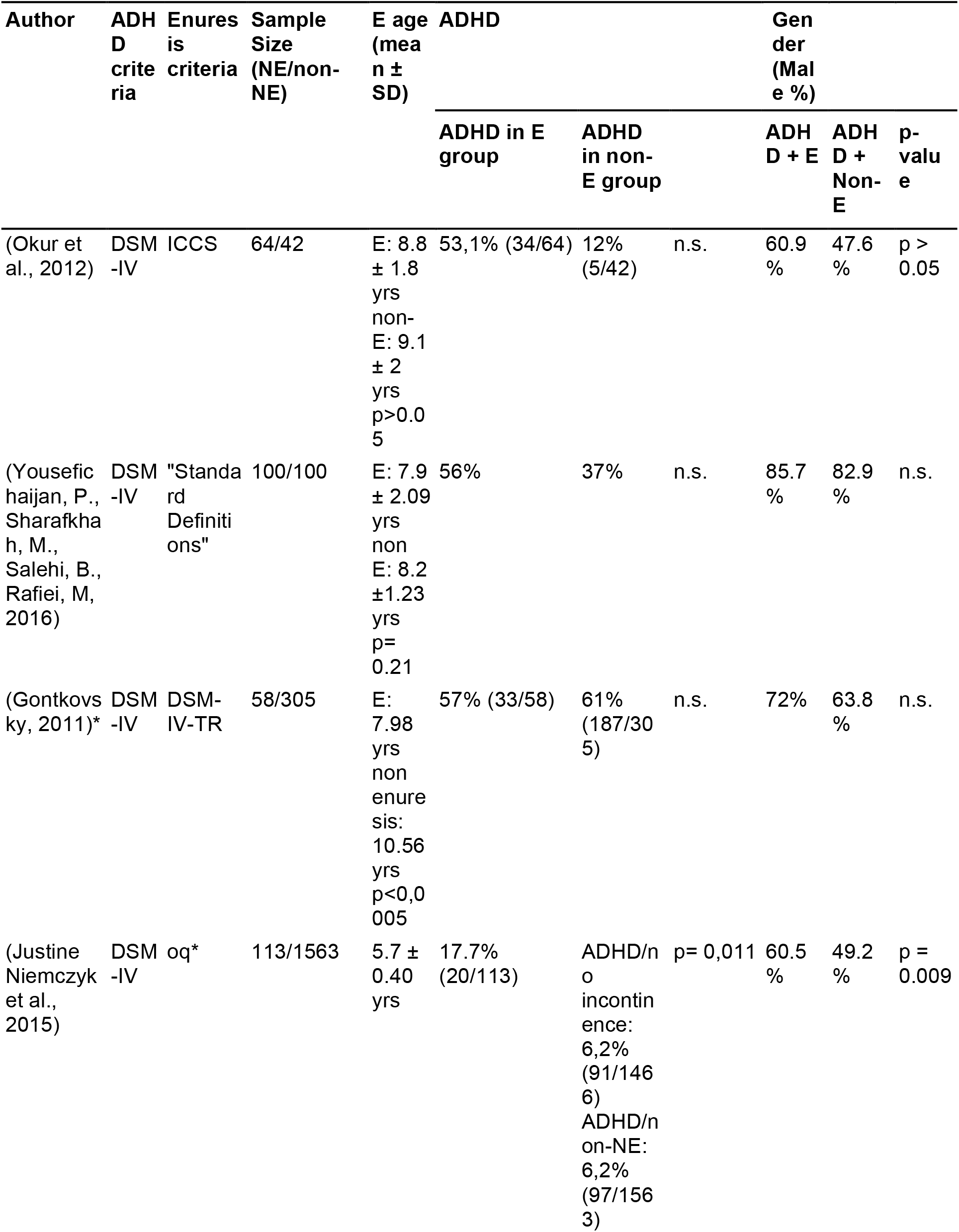
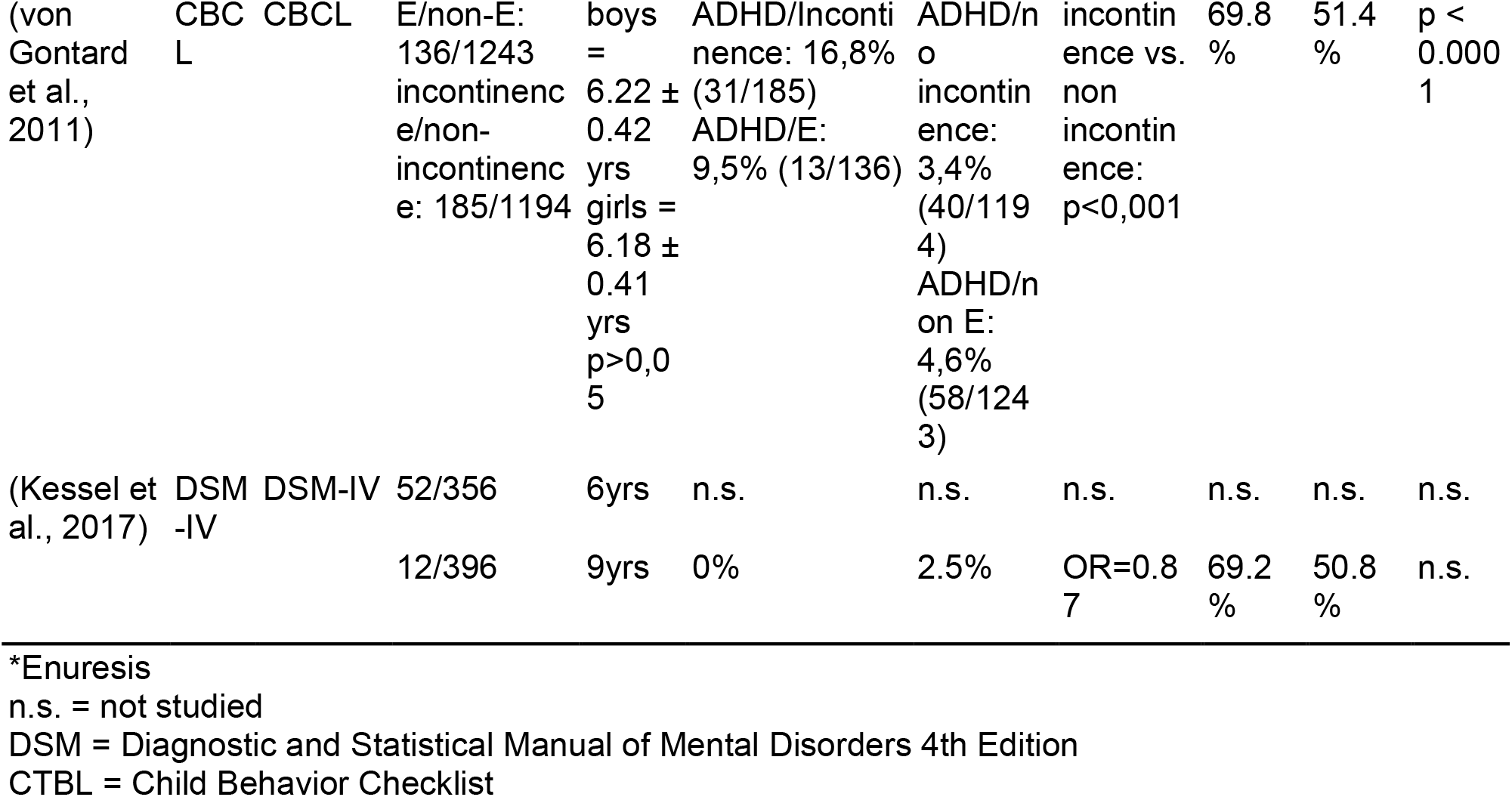
Studies that screened patients with enuresis (E) and without (non-E) for attention deficit hyperactivity disorder (ADHD)

**Table 7.** Enuresis (E) and attention deficit hyperactivity disorder (ADHD) co-occurrence by ADHD subtypes

| Author | Sample Size |  | ADHD subtypes |  |  |  |  |  |
| --- | --- | --- | --- | --- | --- | --- | --- | --- |
|  | E+ADHD | non-E+ADHD | Combined |  | Inattentive |  | Hyperactive–Impulsive |  |
|  |  |  | E | Non-E | E | Non-E | E | Non-E |
| (Elia et al., 2009) | 58 | 286 | 63.8% | 65% | 5.2% | 26.15% | 12% | 8.85% |
| (Yang et al., 2013) | 15 | 38 | 6.7% | 26.3% | 73.3% | 57.9% | 20% | 15.8% |
| (Amiri et al., 2017) | 28 | 132 | 78.6% | 71.2% | 17.9% | 18.2% | 3.5% | 10.6% |
| (Khazaie et al., 2018) | 111 | 220 | 36% | 68.8% | 34.2% | 5% | 29.8% | 32.2% |
| (Ghanizadeh, 2010) | 20 | 151 | 25% | 45% | 45% | 23.8% | 30% | 31.2% |
| (Okur et al., 2012) | 34 | 5 | 23.5% | 20% | 50% | 40% | 26.5% | 40% |
| (Yousefichaijan, P., Sharafkhah, M., Salehi, B., Rafiei, M, 2016) | 56 | 37 | 26.8% | 43.2% | 28.6% | 13.6% | 44.6% | 43.2% |
| (Baeyens, Roeyers, D’Haese, et al., 2007) | 23 (Non-tertiarycare) | - | 21.7% | - | 47.8% | - | 30.5% | - |
|  | 48 (Tertiary care) | - | 37.5% | - | 56.25% | - | 6.25% | - |
| (Van Herzeele et al., 2015) | 8 | - | 25% | - | 62.5% | - | 12.5% | - |
| (Abd-Elmoneim et al., 2020) | 40 | - | 35% | - | 60% | - | 5% | - |
| (Gontkovsky, 2011) | 33* | - | 81.8% | - | 9% | - | 0% | - |
| (Justine Niemczyk et al., 2015) | 10 | - | 20% | - | 20% | - | 60% | - |

\*Three Enuresis+ADHD were "Not Otherwise Specified"

### 4.2) How frequent is ADHD and Enuresis comorbidity?

Frequency of co-occurrence of ADHD and enuresis were addressed through different approaches, as described in 4.1.

Studies that assessed enuresis rates among ADHD children found different results. As shown in Table 3, eight articles were included in this category, being seven cross sectional studies and one case control study. For cross sectional designs, enuresis rates ranged from 10.4% (Ghanizadeh, 2009) to 33.5% (Khazaie et al., 2018). Enuresis rate was higher in the case control study conducted by Zavadenko, et al, who found a rate of 42.7% (Zavadenko et al., 2011).

Frequency of ADHD among children with enuresis appears to be similar. Studies with this approach are displayed in Table 4, being three cross sectional studies, one case control and one cohort study (Baeyens, Roeyers, Van Erdeghem, et al., 2007). In cross sectional studies, ADHD rates ranged from 26% (Van Herzeele et al., 2015) to 74.9% (Amiri et al., 2017). Intermediate values were found by Baeyens D, et al, that compared ADHD rates in children with enuresis in both tertiary-care (40%) and non-tertiary care (29%) (Baeyens, Roeyers, D’Haese, et al., 2007). The case control study (Abd-Elmoneim et al., 2020) also found a frequency of 40% of ADHD among children with enuresis. The cohort study followed enuretic children for four years and its results ranged between 40% and 46% (Baeyens, Roeyers, Van Erdeghem, et al., 2007).

Seven studies that compared enuresis rates in ADHD and non-ADHD children were included, being one cross sectional study, two cohort studies and four case controls. The cross sectional study had the larger sample size (14,900 ADHD/59,600 non ADHD), and found a statistically significant difference between these groups (1,15% vs 0,48%; p<0,001) (Tsai et al., 2017). Data from cohort studies are discussed in section “4.5) Age differences’’. Three out of four case control studies found statistically significant differences between ADHD and non-ADHD groups. Data points are shown in Table 5. Data from these case-control studies were pooled to estimate the enuresis prevalence comparing ADHD and control samples. These studies were less heterogeneous (p = 0.25) or I^2^ lower than 50%, Fixed effect model was used. As shown in Figure 2, the combined OR was 2.49 (95% CI 2.13–2.93, p < 0.001), which indicated that enuresis rates are higher in children with ADHD than in those without this condition (No-ADHD).

**Figure 1.**
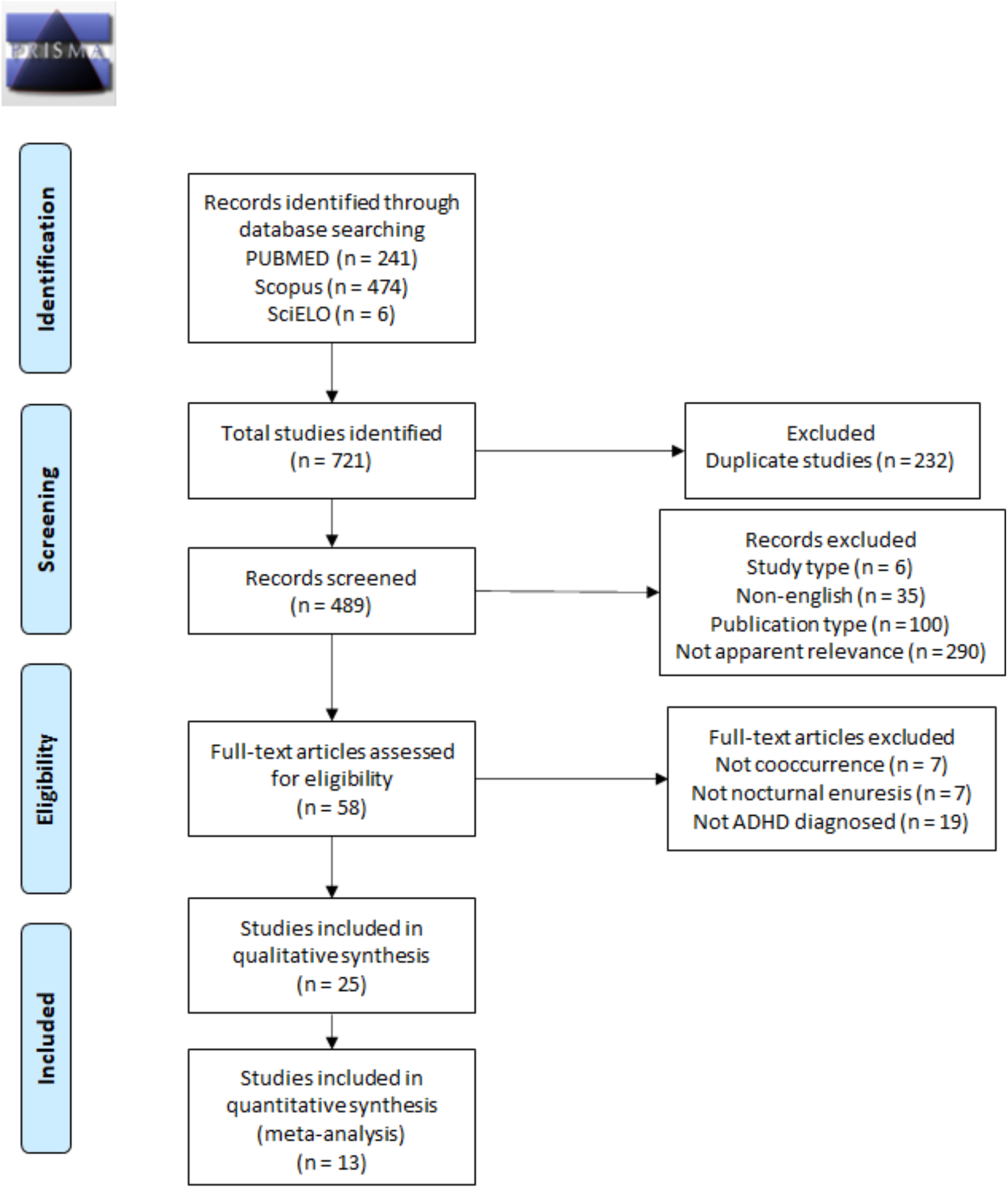
PRISMA flow diagram for studies of ADHD and enuresis co-occurrence.

**Figure 2.**
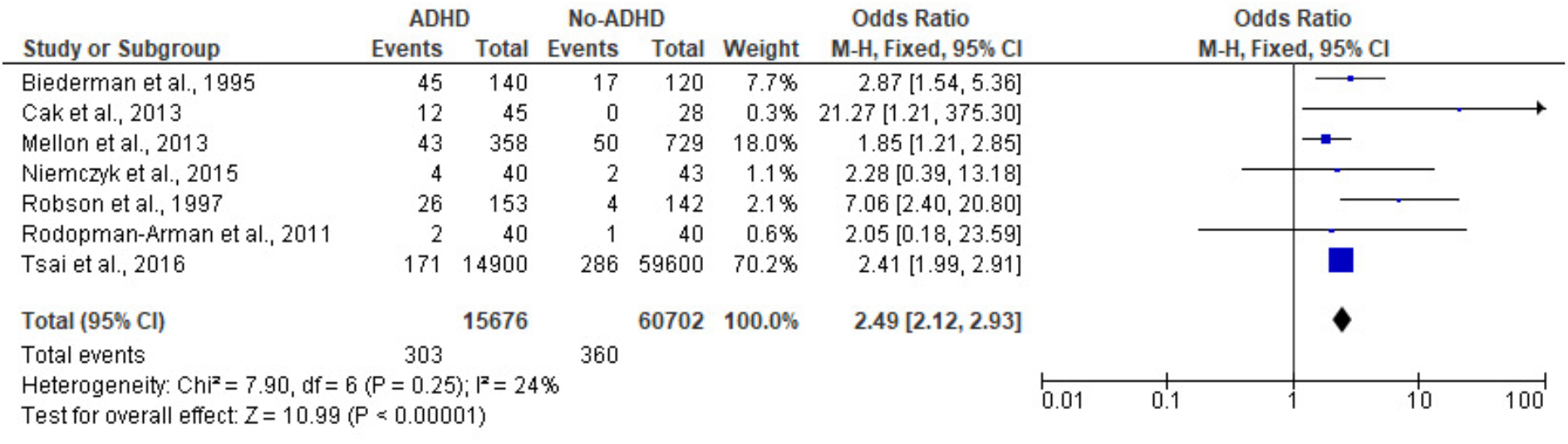
Meta-analysis of enuresis frequencies in ADHD and non-ADHD children.

Finally, six studies - three cross sectionals, two case controls and one cohort - compared ADHD frequencies in children with and without enuresis. Two out of three cross sectional studies found significant differences in ADHD frequency between groups (Justine Niemczyk et al., 2015; von Gontard et al., 2011). For the two case control studies, results were reported segregated by ADHD subtype, but did not provide significant values for ADHD frequency in general, as shown in Table 6 (Okur et al., 2012; Yousefichaijan, P., Sharafkhah, M., Salehi, B., Rafiei, M, 2016; Abd-Elmoneim et al., 2020; Khazaie et al., 2018; Okur et al., 2012). These findings are reported in section “4.3) Association between ADHD types and Enuresis”. Significant values were not found in the cohort study (Kessel et al., 2017). As there is a significant heterogeneity among studies (p = 0.001) and I^2^ higher than 50% (I^2^ = 75%), a Random effect model was used. As shown in Figure 3, the combined OR was 2.03 (95% CI 1.20–3.42, p < 0.001), which indicated that enuresis rates are higher than no-enuresis in ADHD samples.

**Figure 3.**
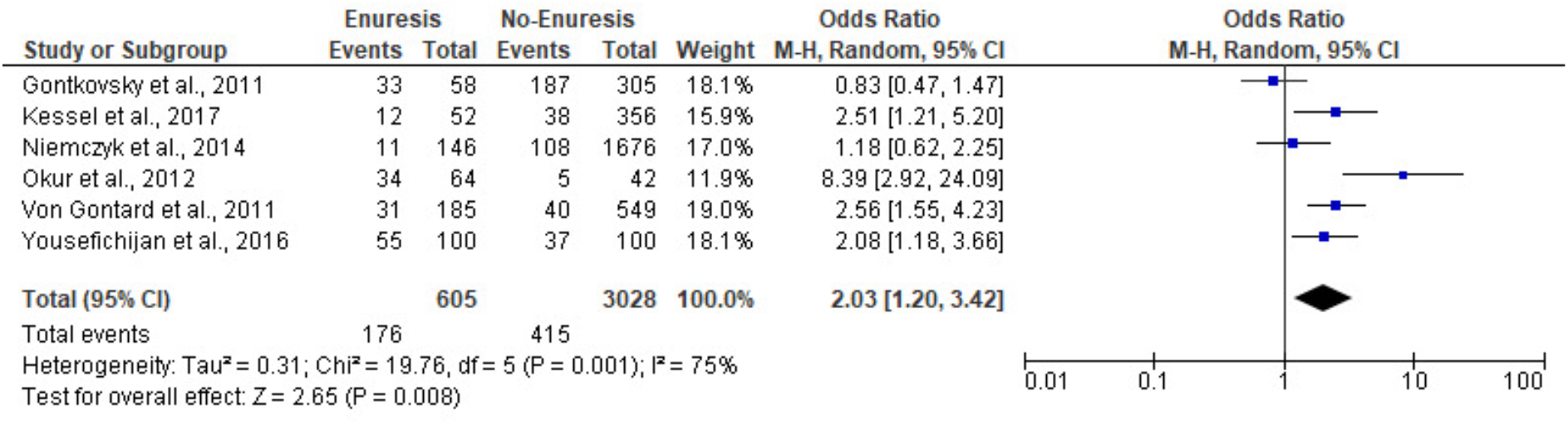
Meta-analysis of ADHD frequencies in children with and without enuresis.

### 4.3) Association between ADHD types and Enuresis

The majority of selected studies did not find or did not address any association between enuresis and ADHD subtype. Only four studies found such association (Abd-Elmoneim et al., 2020; Khazaie et al., 2018; Okur et al., 2012; Yousefichaijan, P., Sharafkhah, M., Salehi, B., Rafiei, M, 2016). A cross sectional study that screened ADHD children for enuresis and found it to be more frequent among ADHD-I children when compared with other subtypes (ADHD-C = 22.5%; ADHD-I = 77.5%; ADHD-HI = 31.7%; p<000.1) (Khazaie et al., 2018). A study with similar design, in spite of not finding significant difference between ADHD subtypes, found that ADHD children with enuresis scored higher in assessment of clinical symptoms of inattention in comparison with non-enuretic ADHD children (p=0.03) (Elia et al., 2009).

Three case-control studies screened children with nocturnal enuresis and those without for ADHD (Abd-Elmoneim et al., 2020; Okur et al., 2012; Yousefichaijan, P., Sharafkhah, M., Salehi, B., Rafiei, M, 2016). One of them showed that ADHD-C and ADHD-I are more frequent in the enuresis group (p=0.04 and p=0.02, respectively) (Okur et al., 2012); and the other found that only ADHD-I was more frequent in the enuretic group (p=0.01) ((Abd-Elmoneim et al., 2020; Khazaie et al., 2018; Okur et al., 2012; Yousefichaijan, P., Sharafkhah, M., Salehi, B., Rafiei, M, 2016)). Finally, another case control study that screened children with enuresis for ADHD found that 60% of the enuretic children had ADHD-I (p<0.001) (Abd-Elmoneim et al., 2020).

### 4.4) Gender differences

The gender difference was assessed in several studies as represented in the summary tables. Among the ones that screened enuresis in ADHD children (Table 3 and Table 5), male were from 57% (Robson et al., 1997) to 88.6% (Yang et al., 2013) of the ADHD patients, (one study reported 100% of male among ADHD, however, the study sample was exclusively composed by male (Biederman et al., 1995). In those studies, when it comes to ADHD and Enuresis co-occurrence group, no significant difference was found, the male rate was between 60.7% (Amiri et al., 2013) to 95% (Ghanizadeh, 2009). However, one study found a significant variation of gender in the ADHD and Enuresis combined group, the male percentage raised from 56.8% in the non-NE group to 72% in the NE group, p-value=0.007 (Khazaie et al., 2018).

Moreover, among the studies that screened ADHD in Enuretic children (Table 4 and Table 6) from 60.5% (J. Niemczyk et al., 2015) to 85.7% (Yousefichaijan, P., Sharafkhah, M., Salehi, B., Rafiei, M, 2016) of the enuretic patients were male; and from 64.7% (Okur et al., 2012) to 88% (Baeyens, Roeyers, Van Erdeghem, et al., 2007) of the ADHD and Enuresis combined group was male. Only one of those studies identified a significant gender variation in the Enuresis and ADHD combined group: the percentage of males in the Enuresis and no ADHD group was 59%, while in the Enuresis and ADHD combined group, the percentage was 88% (p-value=0.01) (Baeyens, Roeyers, Van Erdeghem, et al., 2007).

### 4.5) Age differences

Regarding specifically age differences, many studies did not find or did not show significant results (Amiri et al., 2017; Biederman et al., 1995; Elia et al., 2009; Ghanizadeh, 2009, 2010; Gontkovsky, 2011; J. Niemczyk et al., 2015; Justine Niemczyk et al., 2015; Okur et al., 2012; Rodopman-Arman et al., 2011; Tsai et al., 2017; von Gontard et al., 2011; Yang et al., 2013; Yousefichaijan, P., Sharafkhah, M., Salehi, B., Rafiei, M, 2016). In particular, none of the studies that screened both enuretic and non enuretic children for ADHD, found or mentioned such differences (Amiri et al., 2017; Gontkovsky, 2011; J. Niemczyk et al., 2015; Okur et al., 2012; von Gontard et al., 2011; Yousefichaijan, P., Sharafkhah, M., Salehi, B., Rafiei, M, 2016) One of them, in spite of not reaching significant values, found that the mean age for bladder control was higher in the ADHD group (J. Niemczyk et al., 2015).

Three case controls, one cross sectional study and three cohort studies found significant age differences between the analyzed groups (Abd-Elmoneim et al., 2020; Baeyens, Roeyers, D’Haese, et al., 2007; Baeyens, Roeyers, Van Erdeghem, et al., 2007; Cak et al., 2013; Khazaie et al., 2018; Mellon et al., 2013; Robson et al., 1997).

Two case control studies screened enuretic children for ADHD. One of them was conducted in a tertiary care unit and found that children with both ADHD and enuresis were significantly younger than children with enuresis only (p = 0.009) (Abd-Elmoneim et al., 2020). The other one compared those rates in tertiary and non-tertiary care and found that children in tertiary care were significantly older (p=0.01) and that 9-12 year-olds were more likely to meet criteria for inattention than 6-8 year-olds (48% vs. 31%, p=0.03) (Baeyens, Roeyers, D’Haese, et al., 2007).

The remaining case-control compared ADHD and non-ADHD individuals and found that, at six years old, ADHD children were 2.7 times more likely to present nocturnal enuresis (Robson et al., 1997). The cross sectional study conducted an enuresis screening in an ADHD population and found it to be more frequent among 7-9 year olds and less common in older children (Khazaie et al., 2018).

The included cohort studies addressed the association between spontaneous healing rates and age. A cohort of enuretic children found that the frequency of enuresis in the ADHD group decreased from 65% to 42% within a year, while the frequency of enuresis in the non ADHD group varied from 37% to 39.1% (Baeyens, Roeyers, Van Erdeghem, et al., 2007)). Two studies followed both ADHD and non-ADHD children, screened them for enuresis and reevaluated them in adolescence. In one of them, 26.7% of ADHD children had nocturnal enuresis during childhood, but none of them presented it in adolescence (Cak et al., 2013). Another cohort also reported a dramatic decrease in enuresis rates when comparing results from six and 15 years old in both ADHD and non-ADHD groups. However, at 15 years old, ADHD children were 2.1 times more likely to present enuresis (Mellon et al., 2013).

### 4.6) Comorbidities

Comorbid disorders in ADHD and enuresis combined groups were assessed in eight studies. Only two of them found a significant association between the presence of comorbid disorders and concomitant ADHD and enuresis (Abd-Elmoneim et al., 2020; Tsai et al., 2017; Zavadenko et al., 2011).

Zavadenko, N. N. et al. found higher frequency of comorbidities in patients with ADHD and enuresis than in those with ADHD only (77.4% vs. 60.6%, p < 0.05). Comorbidities included anxiety disorders (54.7% vs. 39.4%, p < 0.05), generalized anxiety disorder (20.8% vs. 12.7%, p < 0.05), obsessive-compulsive disorder (30.2% vs. 22.5%, p < 0.05). The study also assessed different comorbidities according to age groups. Patients at 5 to 9 years old with ADHD and enuresis had a higher frequency of oppositional-defiant behavioral disorder (34.4% vs. 26.7%) and encopresis (9.4% vs. 4.4%), while at the 10–14 years old, obsessive-compulsive disorder (42.9% vs. 23.1%) and tics (14.3% vs. 7.0%) were more common (Zavadenko et al., 2011). It is important to mention that this study did not provide information on cases and controls selection criteria.

Three studies also assessed oppositional-defiant behavioral disorder and no significant difference was found between groups (Elia et al., 2009; Gontkovsky, 2011; Yang et al., 2013). Anxiety disorder was analyzed by two other studies without differences (Biederman et al., 1995; Elia et al., 2009). The same was true to encopresis (Gontkovsky, 2011). Obsessive-compulsive disorder was assessed by one other study with no significant score difference between Enuresis and ADHD combined and ADHD only, however, Enuresis and ADHD group scored higher than Enuresis and ADHD group and Enuresis only group (Abd-Elmoneim et al., 2020).

Tsai, J. D. et al. found that children with enuresis and comorbidity had a greater risk of ADHD than those without nocturnal enuresis and comorbidities (adjusted OR=8.43, 95% CI 4.38 to 16.2). The study considered comorbidities as: epilepsy, mental retardation, autism, Tourette syndrome, sleep disorder, emotional problem, acute reaction to stress, and adjustment disorder (Tsai et al., 2017). Sleep disorder (Yang et al., 2013) and mental retardation was also assessed by other studies that did not find differences between groups (Gontkovsky, 2011).

### 4.7) Socioeconomic factors

Five studies assessed socioeconomic status, social class or parent’s occupation and there was no significant association between these aspects and ADHD and enuresis co-occurrence (Abd-Elmoneim et al., 2020; Biederman et al., 1995; Elia et al., 2009; Ghanizadeh, 2010; Van Herzeele et al., 2015). However, in one of those studies children with Enuresis only, Enuresis and ADHD and ADHD were coming mostly from rural areas and had mainly very low or low socioeconomic status.

## 5) Discussion

Findings reported in topic 4.2, “How frequent is ADHD and Enuresis comorbidity?”, provide evidence of a reciprocal association between ADHD and enuresis occurrence.

Studies that assessed ADHD rates in enuretic children and studies that assessed enuresis in ADHD children found similar results, showing that both conditions are significantly overlapped. The odds ratio obtained in our meta-analysis was two-times higher for both enuresis in ADHD patients as well for ADHD in enuretic children. Many studies have clinical samples and generally the studies with higher co-occurrence rates were poorly evaluated in NOS scale, as shown in Table 1, due to its lack of information on selection of cases and controls (Zavadenko et al., 2011); or were samples taken from tertiary care units (Abd-Elmoneim et al., 2020; Amiri et al., 2013; Baeyens, Roeyers, D’Haese, et al., 2007). Since more complex and severe cases are usually referred to tertiary care units, higher rates of ADHD and enuresis co-occurrence are expected.

Among studies that screened ADHD and non ADHD children for enuresis, enuresis was more frequent among ADHD children. Though, it is important to highlight that the prevalence of enuresis decreased as the sample sizes increased. In spite of that, only one study did not find statistically significant differences between these groups, probably due to its small sample size (40 individuals in the ADHD group) added to the fact that 77,5% was on pharmacotherapy with methylphenidate (J. Niemczyk et al., 2015). Indeed, one of the major limitations of our review is the fact that available literature often did not specify if subjects were on pharmacotherapy or not, and, if they were, did not discuss the effect of treatment on the prevalence of enuresis. Therefore, these data may indicate that the prevalence of enuresis might be underestimated. Further studies on that matter are needed in order to build more robust evidence.

As for the six studies that screened children with and without enuresis for ADHD, only one out of three cross sectional studies did not find significant differences in ADHD frequency between the two groups (Gontkovsky, 2011), what may be due to the fact that the enuresis sample was much smaller than in the other two cross sectional studies: 58 individuals vs. 113 (Justine Niemczyk et al., 2015)and 136 (von Gontard et al., 2011). The latter, in fact, did not conduct a statistical analysis on the differences between the frequency of ADHD among children with and without enuresis, but among children with incontinence problems in general compared with continent children (von Gontard et al., 2011). Two case control studies segregated values according to ADHD subtype, but did not provide statistical analysis regarding differences in ADHD frequency in general (Okur et al., 2012; Yousefichaijan, P., Sharafkhah, M., Salehi, B., Rafiei, M, 2016) Data regarding only enuresis was extracted when possible, as shown in Table 6.

As for our findings regarding the association between ADHD types and occurrence of enuresis, ADHD-C and particularly ADHD-I seem to have a stronger relationship with enuresis, as shown in section 4.3 and Table 8. However, further studies assessing this specific association are required in order to provide more robust evidence, since included studies that addressed the link between enuresis and an specific ADHD subtype had small groups of concomitant ADHD and enuresis, what may bias positive and negative associations (Abd-Elmoneim et al., 2020; Khazaie et al., 2018; Okur et al., 2012; Yousefichaijan, P., Sharafkhah, M., Salehi, B., Rafiei, M, 2016) Besides, samples utilized may not always be representative of the general ADHD population: one of the studies, for instance, included only probands of European descent (Elia et al., 2009).

It is well established that gender plays an important role in ADHD patients and in Enuresis separately. A 2012 systematic review with 163,688 children and adolescents and 14,112 adults found that males were more likely to be diagnosed for ADHD in all ages, the male:female proportion varied from 1.6:1 to 2.4:1 in the overall diagnosis of ADHD; and it was 5.6:1, for ADHD-C in the 13–18-years-old group (Willcutt, 2012). The same is true for enuresis. In a retrospective cross-sectional study with 130,000 children, 7.2% presented NE and 63,7% of those children were male (Ferrara et al., 2020). We found similar results for both conditions in the selected studies. However, in general, studies did not identify a significant gender variation in the Enuresis and ADHD combined group. Consequently, the male prevalence in Enuresis and ADHD combined groups is due to the higher proportion of ADHD and Enuresis alone. Only two studies found a significant variation of gender in the ADHD and Enuresis combined group. One study was retrospective and based on interviews with children and their parents, so the results might be subject to memory bias (Khazaie et al., 2018). The second one was a cohort study, but with a small sample and only 65.8% of follow up rate (Baeyens, Roeyers, Van Erdeghem, et al., 2007).

Enuresis is also known to usually resolve spontaneously in 15% children (Yeung et al., 2006), which is endorsed by our findings. This outcome seems to be also true for ADHD children, since studies that addressed age effects found that the frequency of enuresis decreases as children grow up (Khazaie et al., 2018; Robson et al., 1997). The included studies might be subject to selection and memory bias. The latter is associated with the retrospective study design, that carried out interviews with the children and their parents, and might have overlooked relevant information ((Khazaie et al., 2018). The former is due to the fact that the ADHD sample was taken from a specialty practice, while controls were from a general pediatric population, what may increase frequency differences of enuresis among studied groups (Robson et al., 1997).

Although enuresis seems to heal spontaneously also in ADHD children, it appears to last longer in this group than in non-ADHD individuals. Particularly in tertiary health care units, children were found to be significantly older. The reason for this observation may be attributed to the fact that more complex and resistant cases are usually referred to tertiary centers (Abd-Elmoneim et al., 2020; Baeyens, Roeyers, D’Haese, et al., 2007)

Cohort studies allowed a more precise overview of age effects in concomitant enuresis and ADHD, and support findings of the cross sectional and case control designs (Baeyens, Roeyers, Van Erdeghem, et al., 2007; Cak et al., 2013; Mellon et al., 2013). In spite of that, further cohort studies are required in order to provide more robust evidence, since included studies present limitations such as small samples, heterogeneous control groups and low follow up time (Baeyens, Roeyers, Van Erdeghem, et al., 2007; Cak et al., 2013). One of the reviewed papers, in spite of being a population-based study, was probably not representative of the general population, since the sample included only children from a mainly white, middle class community, which may limit inferences to other populations (Mellon et al., 2013).

In our findings, the screened comorbidities were very heterogeneous between studies. Two studies found a difference between ADHD only or Enuresis only and ADHD and Enuresis combined groups (Tsai et al., 2017; Zavadenko et al., 2011). It is important to mention that Zavadenko et al. study did not provide information on cases and controls selection criteria, so the result may contain selection bias (Zavadenko et al., 2011). In addition, Tsai, J. D. et al. did not collect their original data, once it was obtained through Taiwan’s National Health Insurance Research Database (NHIRD). Thus the authors were not able to check data collect quality and some important information was missing, as clinical conditions, information regarding severity of disorders and seriousness of neuropsychiatric comorbidities (Tsai et al., 2017).

Comorbidities in neurodevelopmental disorders usually are an additive sign of severity of compromise (King et al., 2005). However, in the case of enuresis and ADHD comorbidity, gender distribution, course, age or presence of comorbidities seem not to be different than the isolated conditions. Further studies are necessary to confirm these preliminary findings.

To sum up, our systematic review with meta-analysis supports the higher and reciprocal association between Enuresis and ADHD. The mechanisms beyond the combination of both conditions remain to be elucidated.

## Data Availability

Data will be available upon request.

